# HISTONCHO: A database of intervention histories for onchocerciasis control & elimination in sub-Saharan Africa

**DOI:** 10.1101/2025.05.19.25327763

**Authors:** Matthew A. Dixon, Martin Walker, Aditya Ramani, Jenna E. Coalson, Emily Griswold, Gregory S. Noland, Andrew Tate, Emeka Makata, Ahmed M. A. Ali, Jorge Cano, Paul Bessell, Claudio Fronterrè, Raiha Browning, Wilma A. Stolk, Maria-Gloria Basáñez

## Abstract

In sub-Saharan Africa (SSA), onchocerciasis control has been implemented for many decades, beginning in 1974 under the Onchocerciasis Control Programme in West Africa (OCP) and in 1995 in Central and East Africa (plus Liberia) under the African Programme for Onchocerciasis Control (APOC). Since the establishment of the Expanded Special Project for Elimination of Neglected Tropical Diseases (ESPEN) in 2016, data on mass drug administration (MDA) with ivermectin has been centrally compiled for all endemic countries at implementation unit (IU) level, beginning in 2013. This paper presents HISTONCHO, a database collating detailed information on interventions, including vector control, from 1975 through to 2022, using the ESPEN portal (2013-2022), regional and country reports, implementation partners’ records, and published literature. Reconstructing such intervention histories is crucial for an understanding of their evolution, modelling their impact, and tailoring future interventions. We discuss strengths and limitations associated with the ESPEN database, and how HISTONCHO can be improved to support modelling of intervention strategies as well as onchocerciasis control and elimination efforts by endemic country programmes.

## Background & Summary

Onchocerciasis, known as river blindness, is caused by infection with the filarial nematode *Onchocerca volvulus* and has long posed a considerable public health problem in sub-Saharan Africa (SSA). The parasite is spread among humans though bites from blackfly vector species of the *Simulium* genus that inhabit ecological niches characterised by fast-flowing water bodies (e.g., rivers, rapids).^1^ While currently there are endemic foci in the Arabian Peninsula (Yemen) and Latin America (Brazil and Venezuela), 99% of infected people live in SSA. Infection with *O. volvulus* leads to severe morbidity through skin disease, ocular disease (including irreversible blindness) and epilepsy^2^. It is also associated with excess mortality, particularly in the young with high microfilarial loads^3^. In 2021, the global burden of disease attributable to onchocerciasis was estimated to be 1.26 (0.75–1.90) million disability-adjusted life-years^4^, ranking as causing the 7^th^ highest burden in a list of 21 diseases under the neglected tropical disease (NTD) and malaria category^5^.

Some onchocerciasis foci in SSA have received at least 30 years of mass drug administration (MDA) with ivermectin and up to 31 years of vector control. Despite this, elimination of transmission has been reported only in 8.5% of foci^6^. Interventions in Africa have been delivered by two major regional initiatives, with the Onchocerciasis Control Programme in West Africa (OCP) covering 11 countries,^7^ and the African Programme for Onchocerciasis Control (APOC) expanding to 20 additional countries in Central and East Africa (plus Liberia)^8^. The approximate location of implementation units (IUs) in SSA mapped to different phases of the OCP and APOC countries is shown in Fig. 1.

**Fig. 1.**
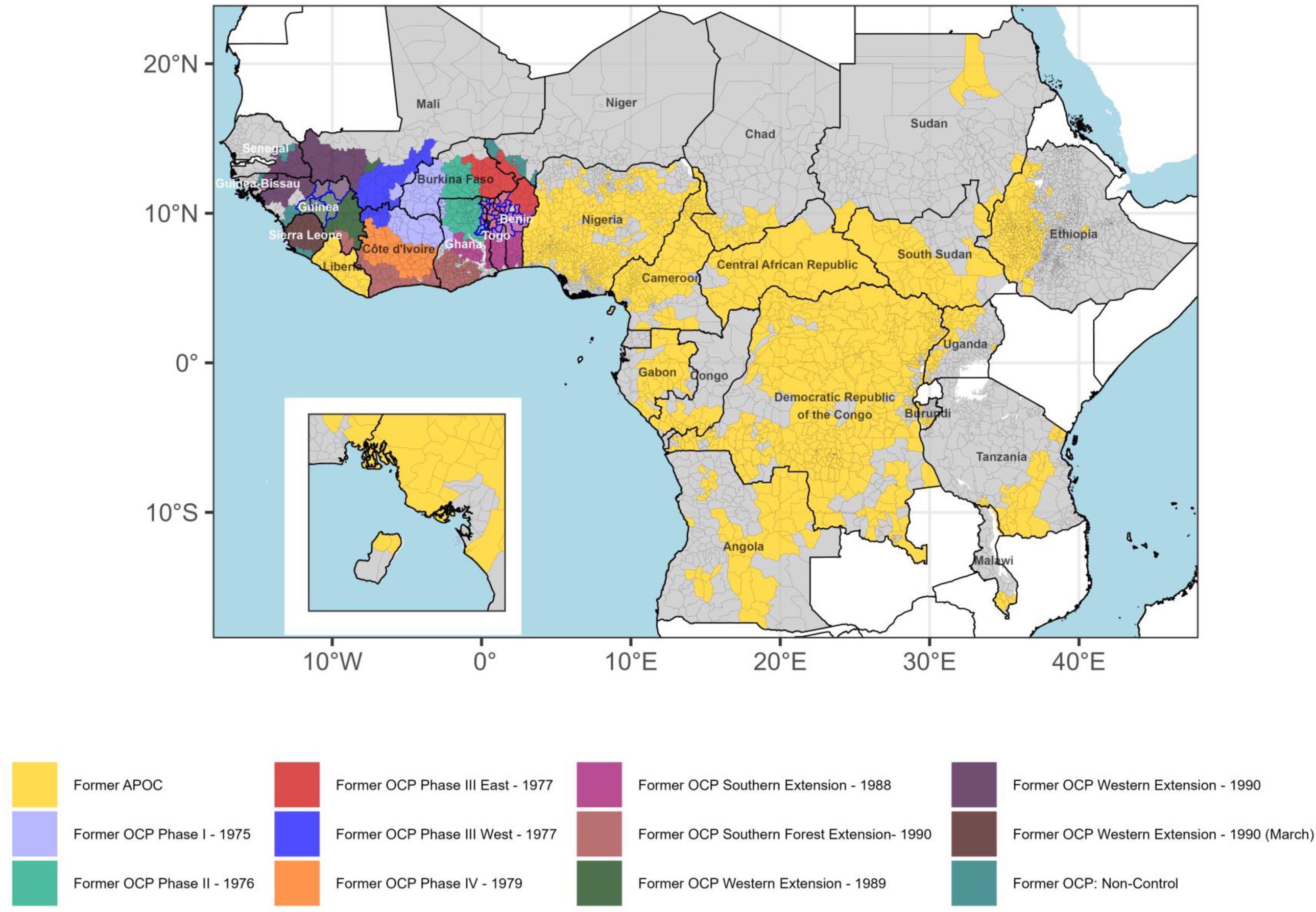
Implementation units (IUs) across sub-Saharan Africa mapped to phases of the Onchocerciasis Control Programme in West Africa (OCP), and the African Programme for Onchocerciasis Control (APOC). IUs are indicated by grey borders. Areas within the OCP identified as non-control, i.e., not included in any of the OCP phases to 2002, are indicated in teal. Special Intervention Zone (SIZ) IUs in former-OCP countries are indicated with blue borders. Inset map shows Bioko Island, Equatorial Guinea. IUs in grey are either non-endemic or with no intervention history (classified as ‘Unknown’ or ‘Not reported’ under the “Endemicity” variable in ESPEN)^22^.

Vector control, based on weekly aerial larviciding of simuliid breeding sites in fast-flowing rivers, over large areas of West Africa, started in the OCP in 1974-1975, targeting areas with the highest blindness prevalence, and aimed at interrupting the transmission of blinding onchocerciasis by implementing interventions for a duration at least as long as that of the lifespan of the adult worm^9^. Initially, the OCP deployed vector control in river basins over a large area across Burkina Faso, Côte d’Ivoire, Ghana, Togo, Benin, Niger and Mali through phases I to IV^9,10^. With the launch of the Mectizan Donation Programme in 1987, ivermectin was made available for mass drug administration (MDA) programmes in the OCP from 1988^11^. Extensions of the original OCP area, using ivermectin MDA, subsequently occurred in parts of Guinea and northern Sierra Leone (western extension from 1989) and in river basins of Côte d’Ivoire, Benin, Ghana, Mali, Senegal and Togo (southern extension from 1988). Vector control was also expanded in these areas to tackle breeding-site locations of re-invading *Simulium damnosum sensu lato* (s.l.)^9^. Ivermectin MDA was further delivered in Guinea Bissau, Mali, Senegal and southern Sierra Leone as part of a western extension to the OCP in 1990^10^. Following the closure of the OCP in 2002, five river basins in West Africa were identified as not having achieved satisfactory entomo-epidemiological indicators^12^. These areas were designated as Special Intervention Zones (SIZ)^13^, located in the Upper Oti river basin in Togo and Ghana, the Upper Ouémé basin in Benin, the Pru basin in Ghana, and both the Upper Niger/Mafou and Tinkisso basins in Guinea. MDA frequency was increased to biannual in the Upper Niger/Mafou and Tinkisso basins, while biannual MDA in conjunction with vector control was implemented in the Upper Oti and Upper Ouémé basins^14^ (Fig. 1). Outside SIZ, the River Gambia/Mako focus in Senegal/Guinea received biannual MDA from 1991^15–17^; the Bougouriba focus in Burkina Faso had 4-monthly MDA (1996-2002); the Rio Corubal focus in Guinea Bissau 3-monthly MDA (1991-1996), and the Rio Géba focus, also in Guinea Bissau, 6-monthly MDA (1989-1996)^17^.

In the remaining onchocerciasis-endemic countries in SSA (beyond the OCP), Rapid Epidemiological Mapping of Onchocerciasis (REMO) was used to identify priority areas for annual ivermectin MDA.^18^ REMO was based on i) the proximity of communities to rivers (and hence likely breeding sites) and ii) the prevalence of palpable onchocercal nodules (which contain adult worms), in samples of adult males, exceeding 20% (an indication that the infection was at least mesoendemic)^18^. These areas were incorporated under APOC from 1995 (Fig. 1). Following the closure of the OCP in 2002, APOC provided technical support to former OCP countries. Large-scale MDA in the late-1980s to mid-1990s relied on mobile teams of drug distributors, a relatively expensive delivery method^7,11^ compared to the Community-Directed Treatment with Ivermectin (CDTI) pioneered by APOC and adopted from the late 1990’s^19^. CDTI aimed to empower communities to deliver MDA, increase coverage, and ensure sustainability^8,11,20^.

APOC closed in 2015, with national programmes continuing MDA under the management of national governments, supported by the Expanded Special Project for Elimination of Neglected Tropical Diseases (ESPEN). The central role of ESPEN has been to provide technical, coordinating and fundraising support to countries across five preventive chemotherapy NTDs, including onchocerciasis^21^. In 2019, ESPEN launched an ambitious data-portal, providing access to subnational programmatic data covering endemicity classifications for each IU and treatment coverage data compiled from 2013 onwards^22^. While this consolidated repository provides an invaluable resource to global health stakeholders, there exists no database detailing onchocerciasis intervention histories at the IU level extending back to the initiation of control across SSA.

Therefore, this paper presents HISTONCHO, a comprehensive database collating, at IU level, intervention histories dating from the start of interventions through merging and enhancing the ESPEN database to support the onchocerciasis control and research communities. An overview of the workflow to identify onchocerciasis-endemic IUs and to compile interventions beginning at the start of control efforts in each IU is illustrated in Fig. 2.

**Fig. 2.**
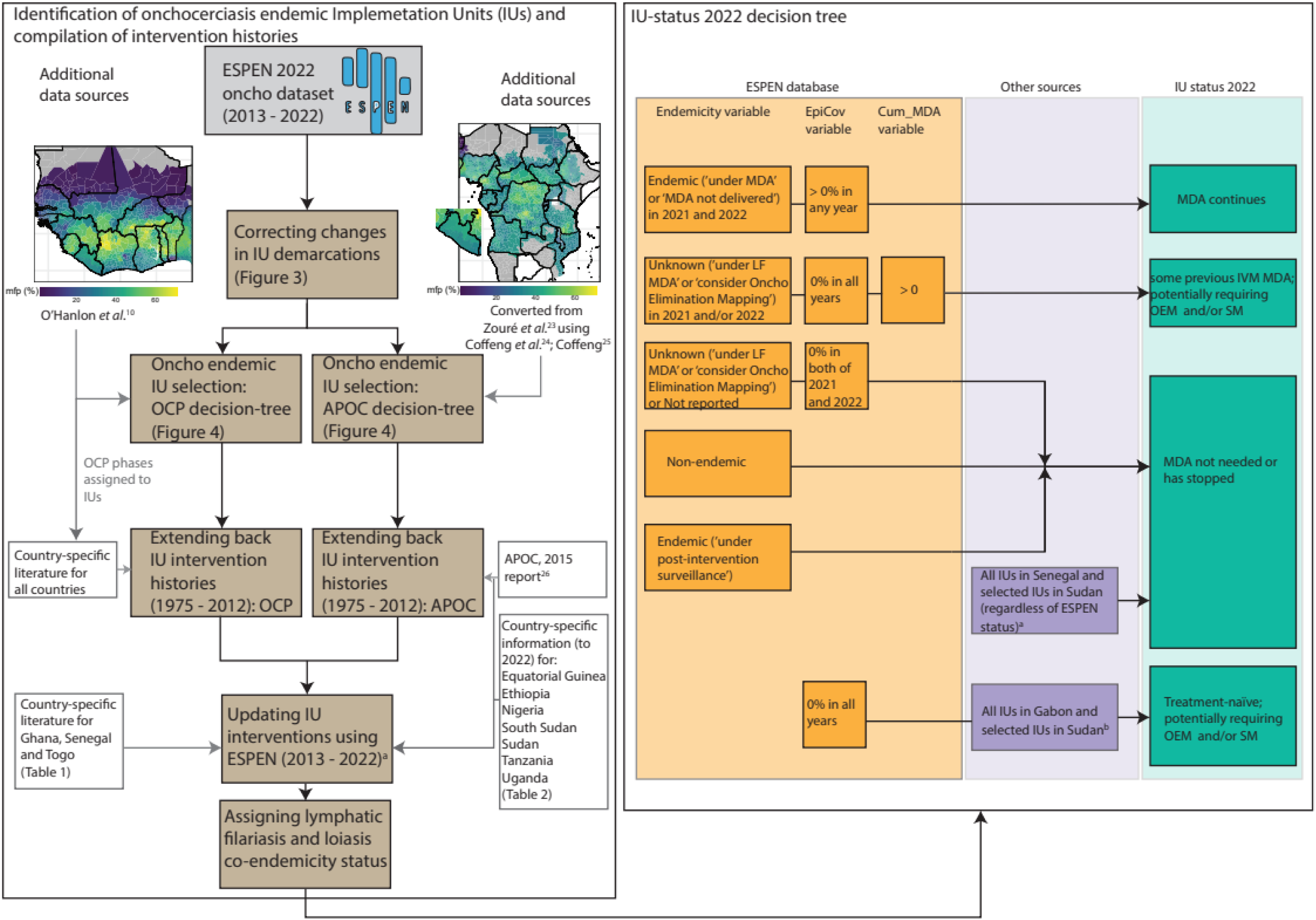
Overview of the workflow to select onchocerciasis-endemic implementation units (IUs) for inclusion to compile intervention histories and define their intervention status in 2022. Entries for former African Programme for Onchocerciasis Control (APOC) countries are taken back to 1975, even though intervention activities only commenced in 1995 (except where country-specific information indicates an earlier start in Nigeria – see Table 2), with entries left-blank for the years prior to the initiation of control. APOC: African Programme for Onchocerciasis Control; ESPEN: Expanded Special Project for Elimination of Neglected Tropical Diseases; IU: Implementation Unit; IVM: ivermectin; LF: lymphatic filariasis; MDA: Mass Drug Administration; mfp: microfilarial prevalence; OEM: Onchocerciasis Elimination Mapping; OCP: Onchocerciasis Control Programme in West Africa; SM: Suitability Mapping. ^a^Indicates that country-specific information (Tables 1 and 2) complements the ESPEN database^22^. ^b^Indicates treatment-naïve status of all IUs in Gabon and 2 IUs Sudan (Table 2).

**Table 1.** Literature sources to inform intervention histories (pre-2013) in former Onchocerciasis Control Programme in West Africa (OCP) countries.

| Country | OCP phase | Mass drug administration (MDA) |  | Vector control |
| --- | --- | --- | --- | --- |
|  |  | Timing/location | Biannual frequency | Timing/location |
| Benin | III-E, SE | O'Hanlon *et al.*<sup>10</sup>; Kale *et al.*<sup>12a</sup>; Abiose *et al.*<sup>14a</sup>; WHO<sup>28a</sup> | Abiose *et al.*<sup>14a</sup> | Kale *et al.*<sup>12a</sup>; Abiose *et al.*<sup>14a</sup>; WHO<sup>28a</sup> |
| Burkina Faso | I, II, III-E | O'Hanlon *et al.*<sup>10</sup> | Borsboom *et al.* (4-monthly)<sup>17</sup> | Koala *et al.*<sup>29</sup> |
| Côte d'Ivoire | I, III-W, IV, SFE | O'Hanlon *et al.*<sup>10</sup>; WHO<sup>30,31</sup>; Koudou *et al.*<sup>32</sup> | – | O'Hanlon *et al.*<sup>10</sup>; WHO<sup>30,31</sup> |
| Ghana | I, II, III-E, SE, SFE | O'Hanlon *et al.*<sup>10</sup>; Kale *et al.*<sup>12a</sup>; Abiose *et al.*<sup>14a</sup>; WHO<sup>33</sup> | Turner *et al.*<sup>34b</sup>; Frempong *et al.*<sup>35</sup>; Biritwum *et al.*<sup>36</sup> | O'Hanlon *et al.*<sup>10</sup>; Kale *et al.*<sup>12a</sup>; WHO<sup>33</sup>; Biritwum *et al.*<sup>36</sup>; Cheke *et al.*<sup>37</sup>; Lamberton *et al.*<sup>38</sup> |
| Guinea | WE-1990, WE-1989, SFE | Hougard *et al.*<sup>9</sup>; O'Hanlon *et al.*<sup>10</sup>; Boatin & Richards<sup>11</sup>; Kale *et al.*<sup>12a</sup>; WHO<sup>39</sup>; Guillet *et al.*<sup>40</sup> | Abiose *et al.*<sup>14a</sup>; Borsboom *et al.*<sup>17</sup> | Hougard *et al.*<sup>9</sup>; O'Hanlon *et al.*<sup>10</sup>; Boatin & Richards<sup>11</sup>; Guillet *et al.*<sup>40</sup> |
| Guinea-Bissau | WE-1989 | Hougard *et al.*<sup>9</sup>; O'Hanlon *et al.*<sup>10</sup>; WHO<sup>41</sup>; Boakye *et al.*<sup>42</sup> | Borsboom *et al.* (3- and 6-monthly)<sup>17</sup> | – |
| Mali | I, III-E, WE-1989, WE-1990 | Hougard *et al.*<sup>9</sup>; O'Hanlon *et al.*<sup>10</sup>; WHO<sup>43</sup>; Dolo *et al.*<sup>44</sup> | – | Koala *et al.*<sup>29</sup>; WHO<sup>43</sup>; Dolo *et al.*<sup>44</sup> |
| Niger | III-E | Hougard *et al.*<sup>9</sup>; O'Hanlon *et al.*<sup>10</sup>; Koala *et al.*<sup>29</sup>; Boakye *et al.*<sup>42</sup>; WHO<sup>45c</sup> | – | – |
| Senegal | WE-1989 | Hougard *et al.*<sup>9</sup>; O'Hanlon *et al.*<sup>10</sup>; Wilson *et al.*<sup>16</sup>; Boakye *et al.*<sup>42</sup>; WHO<sup>45c</sup>; WHO<sup>46,47</sup>; Bagcchi<sup>48c</sup> | Diawara *et al.*<sup>15</sup>; Wilson *et al.*<sup>16</sup>; Borsboom *et al.*<sup>17</sup> | – |
| Sierra Leone<sup>†</sup> | WE-1990 | Kale *et al.*<sup>12a</sup>; Abiose *et al.*<sup>14</sup>; WHO<sup>49</sup>; Kargbo-Labour *et al.*<sup>50</sup> | WHO<sup>49</sup>; Koroma *et al.*<sup>51a</sup> | – |
| Togo | II, III-E, SE | O'Hanlon *et al.*<sup>10</sup>; Kale *et al.*<sup>12a</sup>; WHO<sup>52</sup>; Vinkeles Melchers *et al.*<sup>53d</sup>; Amaral *et al.*<sup>54d</sup> | Vinkeles Melchers *et al.*<sup>53d</sup>; Amaral *et al.*<sup>54d</sup> | Kale *et al.*<sup>12a</sup>; Cheke *et al.*<sup>37</sup>; WHO<sup>52</sup>; Vinkeles Melchers *et al.*<sup>53d</sup>; Amaral *et al.*<sup>54d</sup> |

<sup>a</sup>Location of implementation units (IUs), timing and frequency of interventions under Special Intervention Zones (SIZ) between 2003 – 2012.
<sup>b</sup>All meso- to hyperendemic IUs in Ghana switched to biannual mass drug administration (MDA) in 2009-2010.
<sup>c</sup>Information on (i) Niger submitting a dossier to the World Health Organization (WHO) to verify the elimination of transmission (therefore no further MDA post 2023) and (ii) Senegal applying for WHO verification of elimination after a 3-year post-treatment surveillance (PTS) period, beginning in 2022 across the country (therefore no MDA in IUs from 2022).
<sup>d</sup>Interventions in different regions of Togo, including SIZ period and biannual ivermectin MDA to 2022.
<sup>†</sup>No MDA prior to 2002 assumed due to civil war.

**Table 2.** Literature and data sources used to inform intervention histories in former African Programme for Onchocerciasis Control (APOC) countries.

| Country | Mass drug administration (MDA) |  | Vector control/elimination |
| --- | --- | --- | --- |
|  | Timing/location | Biannual frequency | Timing/location |
| Equatorial Guinea | Herrador *et al.*<sup>55a</sup>; Hernández-González *et al.*<sup>56a</sup> | – | Traoré *et al.*<sup>57a</sup> |
| Ethiopia | FMOH<sup>58b</sup>; R. Retkute (pers. comm.)<sup>b</sup>; Kifle & Nigatu<sup>59c</sup> | FMOH<sup>58b</sup>; R. Retkute (pers. comm.)<sup>b</sup>; Kifle & Nigatu<sup>59c</sup> | – |
| Nigeria | FMOH<sup>60d</sup> | FMOH<sup>61e</sup> and implementation partners (pers. comm.) | – |
| South Sudan | Amaral *et al.*<sup>62</sup> | Colebunders *et al.*<sup>63f</sup> | Jada *et al.*<sup>64f</sup> |
| Sudan | FMOH<sup>65g</sup> and implementation partners | FMOH<sup>65g</sup> and implementation partners; Zarroug *et al.*<sup>66h</sup> | – |
| Tanzania | Mushi<sup>67</sup> | Bhwana *et al.*<sup>68i</sup> | – |
| Uganda | Katabarwa *et al.*<sup>69j</sup> | Katabarwa *et al.*<sup>69j</sup> | Katabarwa *et al.*<sup>69j</sup> |
| Gabon | NA<sup>70</sup> | NA<sup>70</sup> | NA<sup>70</sup> |

## Methods

The main underlying dataset used for selecting onchocerciasis-endemic IUs for inclusion in HISTONCHO was the onchocerciasis ESPEN IU-level data as of 2022^22^, downloaded for each country and combined into a single dataframe.

### Corrections for changes in Implementation Unit (IU) demarcations

During the period covered by the ESPEN data used here (2013-2022), there have been changes to IU boundaries, mostly with some IUs splitting into smaller ones. For example, in 2016, Huambo IU in Angola, was split into two IUs: Cachiungo and Tchikala Tcholohanga, while Huambo continued as a smaller IU within the original larger geographical area (Fig. 3). To create a complete and consistent set of IU histories, we reconciled these subdivisions by synthesising records for the missing years. Specifically, we backfilled data for Cachiungo and Tchikala Tcholohanga using Huambo’s 2013-2015 records. We assumed that the situation in the original IU (Huambo) during those years was representative of the conditions in the derived IUs, including endemicity and MDA status. Fig. 3 illustrates the process to backfill histories in the thus derived IUs, and the rules used to assign their endemicity status.

**Fig 3.**
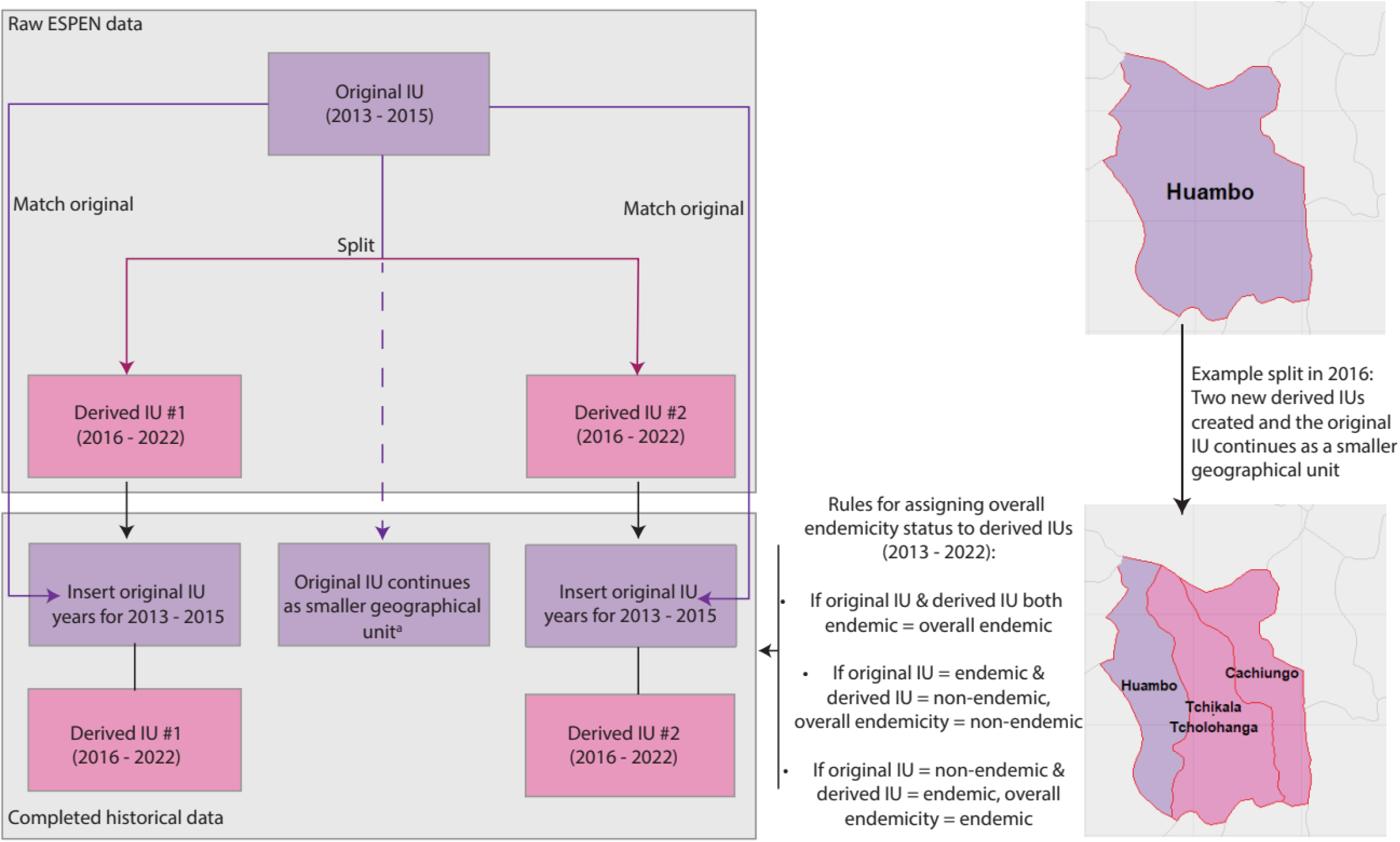
Process for aligning ‘original’ and ‘derived’ implementation units (IUs) in the ESPEN database. In this example, the ‘original’ IU of Huambo, in Angola, was split in 2016 into three smaller units, with two new ‘derived’ IUs plus part of the original, larger IU. ^a^Indicates that the original IU continues as a smaller geographical unit. In other cases, the original IU may have ceased to exist. Within the derived IUs, the original IU intervention history up to the split was used to backfill years from 2013 to the last year prior to the split. Rules for assigning overall endemicity across the derived IUs are described.

### Selecting IUs classified as onchocerciasis endemic with histories of control

IU-level information is available within the ESPEN database not only for IUs classified as endemic, but also for IUs classified as non-endemic—those with ‘Unknown’ endemicity status or ‘Not reported’ status. For former OCP countries in West Africa, the selection criterion for those IUs classified as with ‘Unknown’ or ‘Not reported’ endemicity status was based on whether they were located within the OCP phases shape-files from O’Hanlon *et al*.^10^. In former APOC countries, selection criteria for inclusion of suitable endemic IUs, accounting for those IUs classified as ‘Unknown’ or ‘Not reported’ endemicity status, were based on pre-control prevalence status and whether historical MDA was reported under the “Cum_MDA” variable in the ESPEN database^22^ (which records the cumulative number of MDA rounds for each year). The pre-control prevalence maps used in this process were converted from REMO nodule prevalence maps^23^ to microfilarial prevalence maps using the approach described by Coffeng *et al*.^24^and Coffeng^25^. This method uses a Bayesian hierarchical multivariate logistic regression model to capture the relationship between pre-control nodule prevalence (in samples of adult males aged ≥20 years) and microfilarial (mf) prevalence in the general population (aged ≥5 years)^24^. The model accounts for several key factors, including measurement error in both nodule and mf prevalence, the sensitivity of nodule palpation, and bioclimatic zone (savannah, forest, and forest-savannah mosaic).^24,25^ The mean (point estimate) mf prevalence for an IU was used to assign endemicity classification for each IU (with hypoendemicity corresponding to > 0% but <40% mf prevalence, mesoendemicity to ≥40% but <60%, and hyperendemicity to ≥60% mf prevalence). (The classification of hypoendemicity used here comprises the category of > 0% but <10% referred to as ‘sporadic endemicity’ by O’Hanlon *et al*.^10^ and ≥10% but <40% mf prevalence.) The APOC report was used to confirm the number of onchocerciasis-endemic countries under its umbrella^26^.

Details of each selection step for both former OCP and APOC countries, including the number of IUs included at each step, are illustrated schematically as a decision tree in Fig. 4.

**Fig 4.**
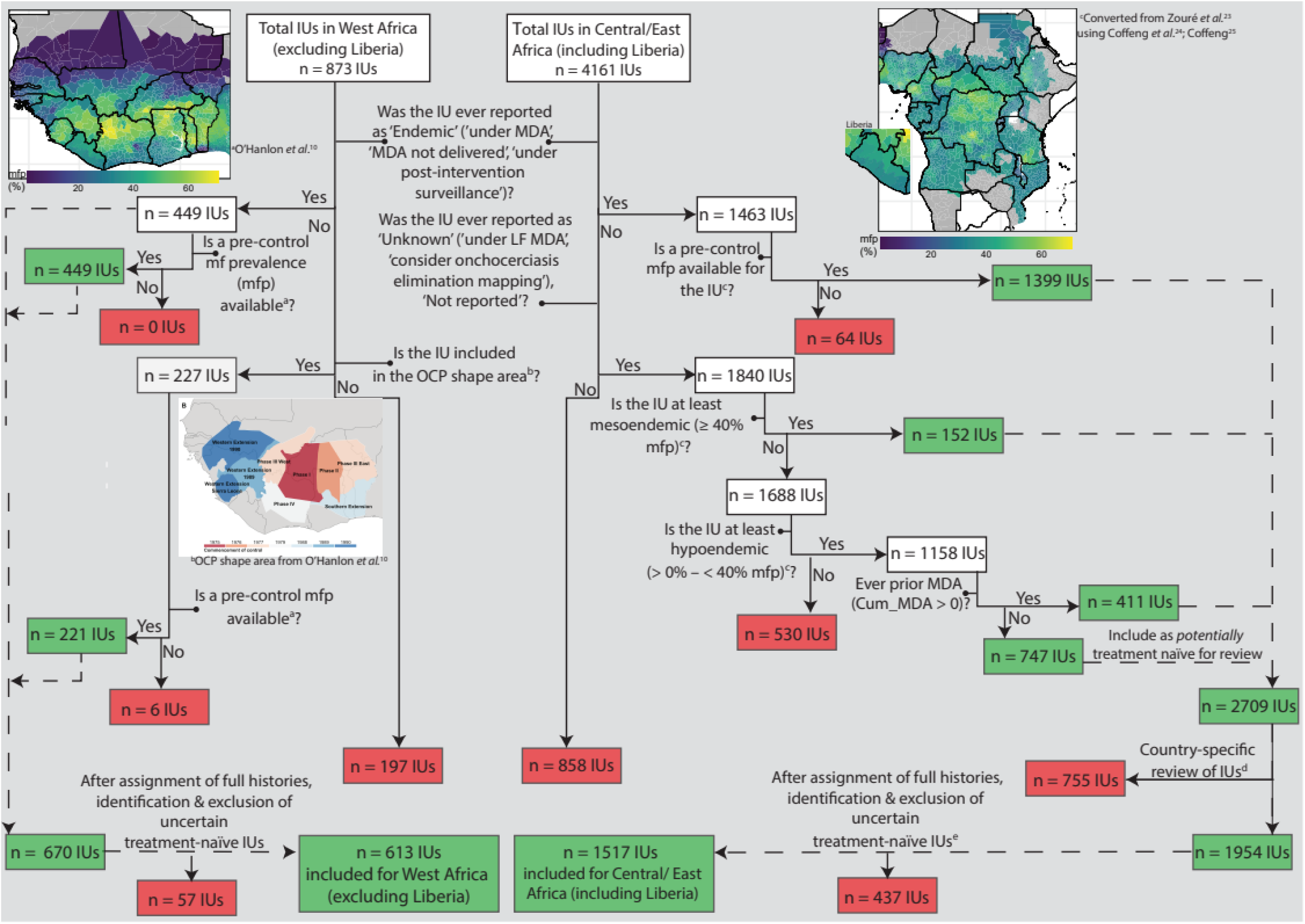
Decision tree to identify onchocerciasis-endemic implementation units (IUs) with intervention histories for former Onchocerciasis Control Programme in West Africa (OCP) and African Programme for Onchocerciasis Control (APOC) countries. Maps show IU-level baseline microfilarial prevalence (mfp) from ^a^O’Hanlon *et al.*^10^ (^b^OCP boundaries) and ^c^converted from nodule prevalence in Zouré *et al.*^23^ to mfp following Coffeng *et al*.^24^ and Coffeng^,25^. ^d^Country-specific review for Ethiopia, Nigeria, Sudan and Tanzania (Table 2). ^e^Excepting 27 IUs in Gabon and 2IUs in Sudan that are clearly documented as treatment-naive^65,70^. Green and red boxes indicate IU inclusion and exclusion, respectively.

### Pre-ESPEN data sources (1975–2012)

Intervention histories for those IUs included as endemic through the decision tree presented in Fig. 4 were extended backwards (backfilled) to 1975 so that the earliest possible start year for interventions could be included prior to ESPEN. Data to inform pre-ESPEN intervention histories on MDA (including timing and location, frequency and total population coverage) and vector control were sought from a range of sources as described below.

In the former OCP countries, shape-files from the map in O’Hanlon *et al.*^10^ (inset map in Fig. 4) were used to map the location of IUs within each country under the different phases of the OCP, which varied in the year of start/end of vector control and start of ivermectin MDA. Additional data sources were identified to inform the start/end and frequency of ivermectin MDA during the OCP and post-OCP (SIZ) until 2012, and to specify the location and features of intervention in the SIZ for each country where appropriate. Table 1 provides a summary of literature and data sources used for the OCP. Treatment coverage was assigned to each IU at 52% of total population prior to 1996, and at 65% of total population (80% of eligible population) from 1996 to 2012. This represents a 25% increase in coverage following the implementation of CDTI (Supplementary File 1, Fig. S1). However, in some areas, the transition to CDTI resulted in an initial decrease of coverage^27^.

In the former APOC countries, MDA coverage was assigned using country-level coverage estimates from the 2015 APOC report^26^. Every onchocerciasis-endemic IU within each former APOC country was assigned a total population coverage of 25% to reflect a (conservative) nominal coverage where the report indicated coverage being less than 65%, or 65% where the reported national coverage was ≥65%. In the HISTONCHO database (see Supplementary File 1, Table S1), raw coverage values (under the variable “Cov_Raw”) are also provided (informed by ESPEN^22^, APOC^26^, and sources cited in Table 2). Specific data sources were used from the start of APOC control programmes until 2022 for Bioko Island in Equatorial Guinea^55–57^, Ethiopia^58,59^, Nigeria^60,61^, South Sudan^62–64^, Sudan^65,66^, Tanzania^67,68^, and Uganda^69^, and for the timing/location of biannual MDA and vector control/elimination where appropriate. Table 2 provides a summary of literature and data sources used for APOC intervention histories.

The updated dataset extracted from ESPEN^22^, including back-dated intervention histories, was subsequently modified to include variables for each IU by year to indicate whether “Non-CDTI_MDA” (i.e., MDA occurring prior to CDTI), “CDTI_MDA”, or “Biannual_CDTI_MDA” were implemented (using a value of 1 where present), “Number_Rounds” (1 or 2 depending on annual or biannual frequency) and total population coverage (“Cov_Raw” for raw coverage and “Cov_Cat” for categorised values). For the 2013-2022 period, “Cov_Raw” corresponds to the “EpiCov” variable in ESPEN with the exception of Ethiopia, Nigeria, Sudan and Uganda (Table 2). A new variable named “Cum_MDA_Rev”, which revises the cumulative MDA variable (“Cum_MDA”) in the ESPEN database^21^, was also included to track the cumulative number of MDA rounds for each IU based on our back-dated histories using the data sources described in this section. A variable to indicate, for each year, the presence of “Vector_Control” was also included, with a 0 for no vector control, 1 for the presence of vector control, or 2 indicating that vector elimination (or disappearance) had taken place.

### ESPEN database (2013-2022) and status of MDA in 2022

Information on MDA coverage at the IU level was extracted directly from ESPEN^22^, covering the years 2013-2022 (version received on 03/October/2023), reflecting updated survey data through to 2022). The ESPEN “EpiCov” variable (defined as epidemiological coverage of total population, and calculated as ‘total treated/reported total population requiring treatment’ from the World Health Organization’s (WHO) Joint Reporting Form (JRF) system^22^) was used to specify (in “Cov_Cat”) a categorical coverage value of 25% (0% < “EpiCov” <65%) or 65% (“EpiCov” ≥65%) for each year. Coverage was set as a categorical variable to account for “EpiCov” values exceeding 100%, and the generally acknowledged uncertainty in reported coverage^71^. As shown in Fig. 2, Table 1 and Table 2, other literature and data sources were also used to compile the histories through to 2022 in certain situations. For example, histories for all years across the whole of Nigeria^60,61^, Sudan^65^ and Uganda^69^ were compiled using sources that contained more detailed information compared to ESPEN, with specific data available on biannual MDA. (ESPEN does not currently include data on the frequency of MDA in IUs.) In certain subnational locations, including the foci of Mahenge and Malinyi in Tanzania^68^, Maridi in South Sudan^63^ and Abu Hamed in Sudan^66^, literature sources provided information on biannual treatment specifically in these areas. Literature on Bioko Island in Equatorial Guinea^55–57^ and Uganda^69^ also provided information on vector control and elimination.

In the ESPEN database^22^, data are very sparse for the first year in 2013. Therefore, where there was no previous MDA indicated by ESPEN (i.e., “Cum_MDA” variable equal to 0 in 2013), and there was no previous MDA included as described above (see **Pre-ESPEN data sources (1975 – 2012)**, no MDA was included for this year. Where there was previous MDA, we assumed that there was MDA in 2013, irrespective of the indication in the ESPEN database^22^. When detailed subnational data were obtained (Table 2), these data were used to inform treatment in 2013.

Defining the IU status in 2022 would permit the use of transmission dynamics modelling to project the impact of future continuation of current interventions or of the adoption of alternative strategies, and assist decision-making by country programmes for onchocerciasis control and elimination. Therefore, a combination of sources was used to inform a variable named “Trt_Status_2022”, with the right-hand side of Fig. 2 outlining the criteria for defining an IU as either ‘MDA continues’,’ ‘MDA stopped’, ‘Treatment-naïve; potentially requiring onchocerciasis elimination mapping (OEM) and/or suitability mapping (SM)’. OEM is based on desk reviews, vector breeding-site assessments and epidemiological surveys using anti-Ov16 serology in adults aged ≥20 years, targeting first-line villages and, if necessary, conducting risk-stratified secondary surveys^72^. Suitability mapping refers to the modelling approach presented by Cromwell *et al*.^73^ which predicts onchocerciasis suitability based on ground-truth data (occurrence of geo-located onchocerciasis cases diagnosed by any parasitological, clinical or serological diagnostic) and using environmental covariates, including climatic factors, vegetation, breeding-site information and urbanicity data.

In addition to the three categories of ‘MDA continues’, ‘MDA stopped’ and ‘Treatment-naïve’, the category of ‘some previous IVM MDA; potentially requiring OEM and/or SM’ was included to indicate cases in ESPEN for which “EpiCov” values are 0, yet “Cum_MDA” values are > 0. The category of ‘some previous IVM MDA; potentially requiring OEM and/or SM’ was further differentiated with an asterisk (*) to indicate IUs in former OCP countries and those without it (not in the former OCP area). Additional data sources reported that interventions have ceased in Senegal—with the country having entered a three-year period of post-treatment surveillance (PTS) since 2022^45,48^—and that no IUs are currently under MDA in Sudan^65^. Therefore, all IUs within these two countries were designated as ‘MDA stopped’. In Gabon, a country with wide-spread co-endemicity of onchocerciasis and loiasis (see **Co-endemicity with lymphatic filariasis and loiasis**), all IUs were designated as ‘Treatment-naïve; potentially requiring onchocerciasis elimination mapping (OEM) and/or suitability mapping (SM)’^70,72,73^.

A variable named “MDA_VC_Mapping” was introduced to summarise the type of intervention history (through to 2022) in each IU. This variable can take the following categorical values: ‘Annual MDA’ where only annual ivermectin MDA has been implemented; ‘Biannual MDA’ if there has been any biannual MDA; ‘Vector control’ if any anti-vectorial measures have been implemented; ‘Annual MDA and vector control’, and ‘Biannual MDA and vector control’ (Supplementary File 1, Table S1). Although biannual MDA was introduced in the River Gambia focus of Senegal since 1991^15–17,27^, it was not possible to extrapolate this (river-basin) localised strategy to the whole of the IU where this focus was located. The same applies to the Mako focus in Guinea, and the Rio Corubal and Rio Géba foci in Guinea Bissau, where more frequent (than annual) treatment was implemented for some years^17^.

### Co-endemicity with lymphatic filariasis and loiasis

Information on co-endemicity with two other vector-borne filarial infections, lymphatic filariasis (LF, caused in Africa by infection with *Wuchereria bancrofti* and transmitted by mosquitoes, also known as elephantiasis), and loiasis (caused by *Loa loa* and transmitted by tabanid flies, also known as African eye worm) was integrated into HISTONCHO. These two filariases are important in the context of onchocerciasis control. In the case of LF, ivermectin is used as part of combination therapy (with albendazole) known as IA (also referred to as ALB-IVM in ESPEN), and LF programmes are often integrated with those for onchocerciasis^6,74^. In the case of *L. loa*, onchocerciasis-loiasis co-endemic areas pose challenges to onchocerciasis control due to the risk of potentially fatal severe adverse events (SAEs) following treatment with ivermectin of individuals with high *L. loa* mf loads^75,76^. Fears associated with present or past SAEs can lead to reduced treatment adherence, which might explain persistence of onchocerciasis in some co-endemic areas with long histories of MDA^77^.

The ESPEN databases for LF^78^ and loiasis^79^ were extracted, and corrections for IU demarcations were made as described above for onchocerciasis (see **Corrections for changes in Implementation Unit (IU) demarcations**). The resulting LF and loiasis databases were compared with the database for onchocerciasis based on IU code and calendar year to inform co-endemicity status^80,81^.

An IU was considered co-endemic with LF, if for any of the years analysed (2013-2021 for LF), endemicity in the ESPEN LF database^78^ was recorded as: ‘Endemic (under MDA)’, ‘Endemic (MDA not delivered)’, ‘Endemic (under post-intervention surveillance)’, or ‘Endemic (pending IA)’. An IU was considered not co-endemic with LF if in the ESPEN LF database^78^ it was classified as ‘Non-endemic’ in at least one year, or ‘Unknown’ (if it was never reported to be either endemic or non-endemic). An IU was considered co-endemic with loiasis if for any of the years analysed (2013-2021 for *Loa*)^79^, endemicity was recorded as ‘Hypo-endemic, ‘Meso-endemic’ or ‘Hyper-endemic’ for loiasis (according to the prevalence of history of eye-worm passage following Rapid Assessment Procedure for Loiasis, RAPLOA)^82,83^. Remaining IUs were categorized as ‘Non-endemic’ or ‘Unknown’ as described for LF.

### Data availability and updates to intervention histories

As new information, through further country-engagement or identification of relevant literature, becomes available, the histories can be updated. HISTONCHO (as RDS and CSV files) is available at the following Zenodo link: https://zenodo.org/records/15390119^84^.

## Data Records

In the HISTONCHO database^84^, 2,130 IUs (Fig. 4) are included as pre-control endemic (with histories of control or well-documented treatment-naïve^65,70^) across 28 countries (613 IUs in former OCP countries and 1,517 IUs in former APOC countries). The 2,130 IUs were distributed across 11 former OCP countries, namely, Benin (n = 70 IUs, 3.3% of all IUs), Burkina Faso (64 IUs, 3.0%), Côte d’Ivoire (106 IUs, 5.0%), Ghana (205 IUs, 9.6%), Guinea (26 IUs, 1.2%), Guinea Bissau (33 IUs, 1.5%), Mali (37 IUs, 1.7%), Niger (14 IUs, 0.7%), Senegal (8 IUs, 0.4%), Sierra Leone (14 IUs, 0.7%) and Togo (36 IUs, 1.7%), and 17 former APOC countries, namely, Angola (49 IUs; 2.3%), Burundi (13 IUs; 0.6%), Cameroon (155 IUs, 7.3%), Central African Republic (29 IUs, 1.4%), Chad (46 IUs, 2.2%), Democratic Republic of the Congo (359 IUs, 16.9%), Ethiopia (219 IUs, 10.3%), Equatorial Guinea (2 IUs, 0.09%), Gabon (27 IUs, 1.3%), Liberia (15 IUs, 0.7%), Malawi (8 IUs, 0.4%), Nigeria (449 IUs, 21.1%), Republic of Congo (17 IUs, 0.8%), Sudan (8 IUs, 0.4%), South Sudan (48 IUs, 2.3%), Tanzania (28 IUs, 1.3%), and Uganda (45 IUs, 2.1%). Of the 2,130 IUs, 1,788 (83.9%) have been under annual MDA, with 272 of these (15.2%) also having had vector control. A total of 313 IUs (14.7%) have received biannual MDA during their intervention history, with 146 of these (46.6%) also having had vector control. The remaining 29 IUs (1.4%), located in Gabon (27 IUs) and Sudan (2 IUs), have not received any intervention (Fig. 5).

**Fig. 5.**
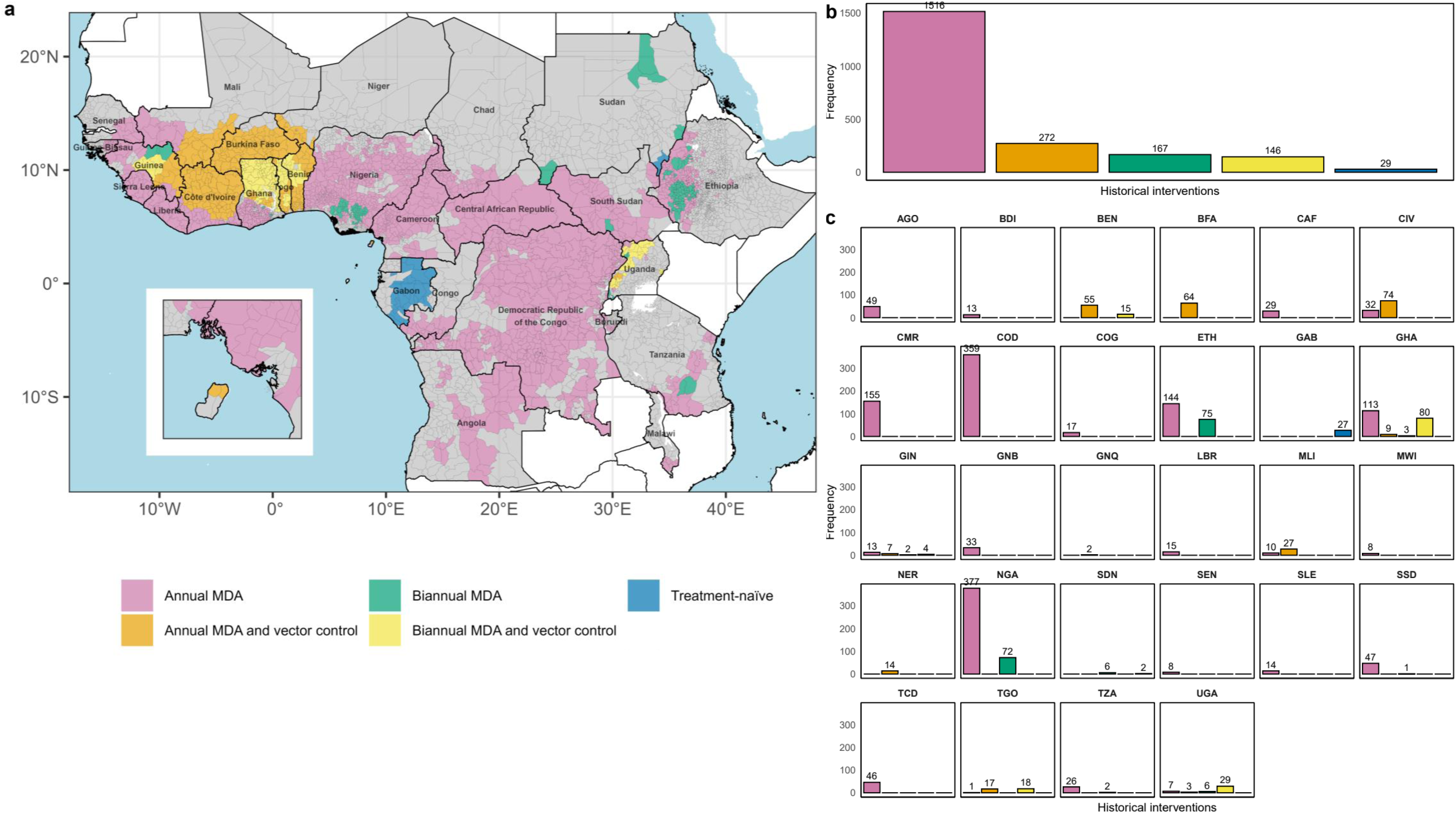
Distribution of historical interventions by implementation unit (IU). Classification of historical interventions is presented: (**a**) across a map of endemic countries, (**b**) as a frequency distribution across all IUs, and (**c**) as a frequency distribution of IUs within each country. IUs are indicated by grey borders. Inset map shows Bioko Island (Equatorial Guinea) in (**a**). Annual and biannual MDA refer to the frequency (yearly or 6-monthly) of ivermectin mass drug administration (MDA). IUs in grey are either non-endemic or with no intervention history (classified as ‘Unknown’ or ‘Not reported’ under the “Endemicity” variable in ESPEN^22^). AGO: Angola; BDI: Burundi; BEN: Benin; BFA: Burkina Faso; CAF: Central African Republic; CIV: Côte d’Ivoire; CMR: Cameroon; COD: Democratic Republic of the Congo; COG: Republic of Congo; ETH: Ethiopia; GAB: Gabon; GHA: Ghana; GIN: Guinea; GNB: Guinea Bissau; GNQ: Equatorial Guinea; LBR: Liberia; MLI: Mali; MWI: Malawi; NER: Niger; NGA: Nigeria; SDN: Sudan; SEN: Senegal; SLE: Sierra Leone; SSD: South Sudan; TCD: Chad; TGO: Togo; TZA: Tanzania; UGA: Uganda. The foci of River Gambia/Mako (Senegal/Guinea) and Rio Corubal/Rio Géba (Guinea Bissau)—with no vector control—had more frequent treatment for some periods^17^ (not mapped to IU-level).

Fig. 6 highlights that the countries with the largest number of MDA rounds are generally concentrated in West Africa, in former OCP countries, where up to 52 rounds of MDA have been deployed in Togo (in 13 IUs), followed by Benin (up to 45 in 15 IUs) and Ghana (up to 4 in 9 IUs). In former APOC countries, the largest number of MDA rounds have been documented in Uganda with up to 40 rounds, followed by Nigeria (35) and Ethiopia (31). The large number of MDA rounds in these countries can be explained by the implementation of biannual MDA and the fact that these countries are not co-endemic with loiasis (Ethiopia, Ghana, Uganda) or if they are, loiasis is mostly hypoendemic (Benin, Nigeria)^81,85^. (No RAPLOA surveys have been conducted in Benin^80^.)

**Fig. 6.**
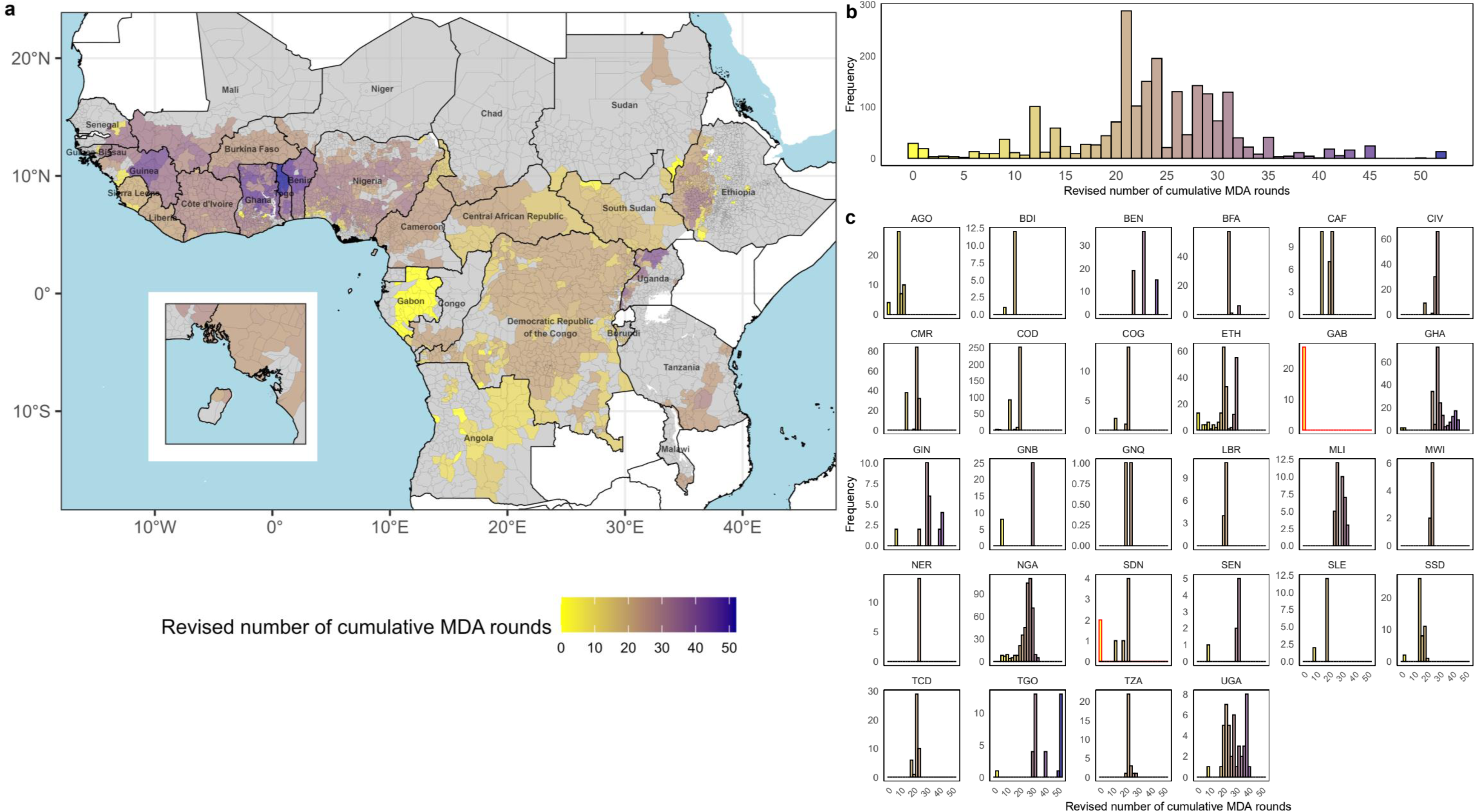
Distribution of revised cumulative number of ivermectin mass drug administration (MDA) rounds by implementation unit (IU). Revised cumulative number of MDA rounds is presented: (**a**) across a map of endemic countries, (**b**) as a frequency distribution across all IUs, and (**c**) as a frequency distribution of IUs within each country. IUs are indicated by grey borders. Inset map shows Bioko Island (Equatorial Guinea) in (**a**). In (**c**), bars with red borders indicate that the revised number of cumulative MDA rounds, “Cum_MDA_Rev” in HISTONCHO^84^ = 0, representing treatment-naïve IUs. IUs in grey are either non-endemic or with no intervention history (classified as ‘Unknown’ or ‘Not reported’ under the “Endemicity” variable in ESPEN)^22^. Country abbreviations are as in Fig. 5. The foci of River Gambia/Mako (Senegal/Guinea) and Rio Corubal/Rio Géba (Guinea Bissau)—with no vector control—had more frequent treatment for some periods^17^ (not mapped to IU-level).

Using the co-endemicity categorisations, 1,426 of the 2,130 IUs are co-endemic with LF (66.9%), 950 are co-endemic with loiasis (44.6%), and 667 are co-endemic with LF and loiasis (31.3%). Co-endemicity with loiasis is mostly concentrated in central Africa (Fig. 7a). Of the 950 IUs that are co-endemic with loiasis, 221 (23.3%) are hyperendemic for loiasis (Fig. 7b). Endemicity categories according to RAPLOA are defined as: no risk (0% to <5%); hypoendemic (low risk) (5% to <20%); mesoendemic (moderate risk) (20% to <40%), and hyperendemic (high risk) (≥ 40%) prevalence of eye-worm history^79,80^.

**Fig. 7.**
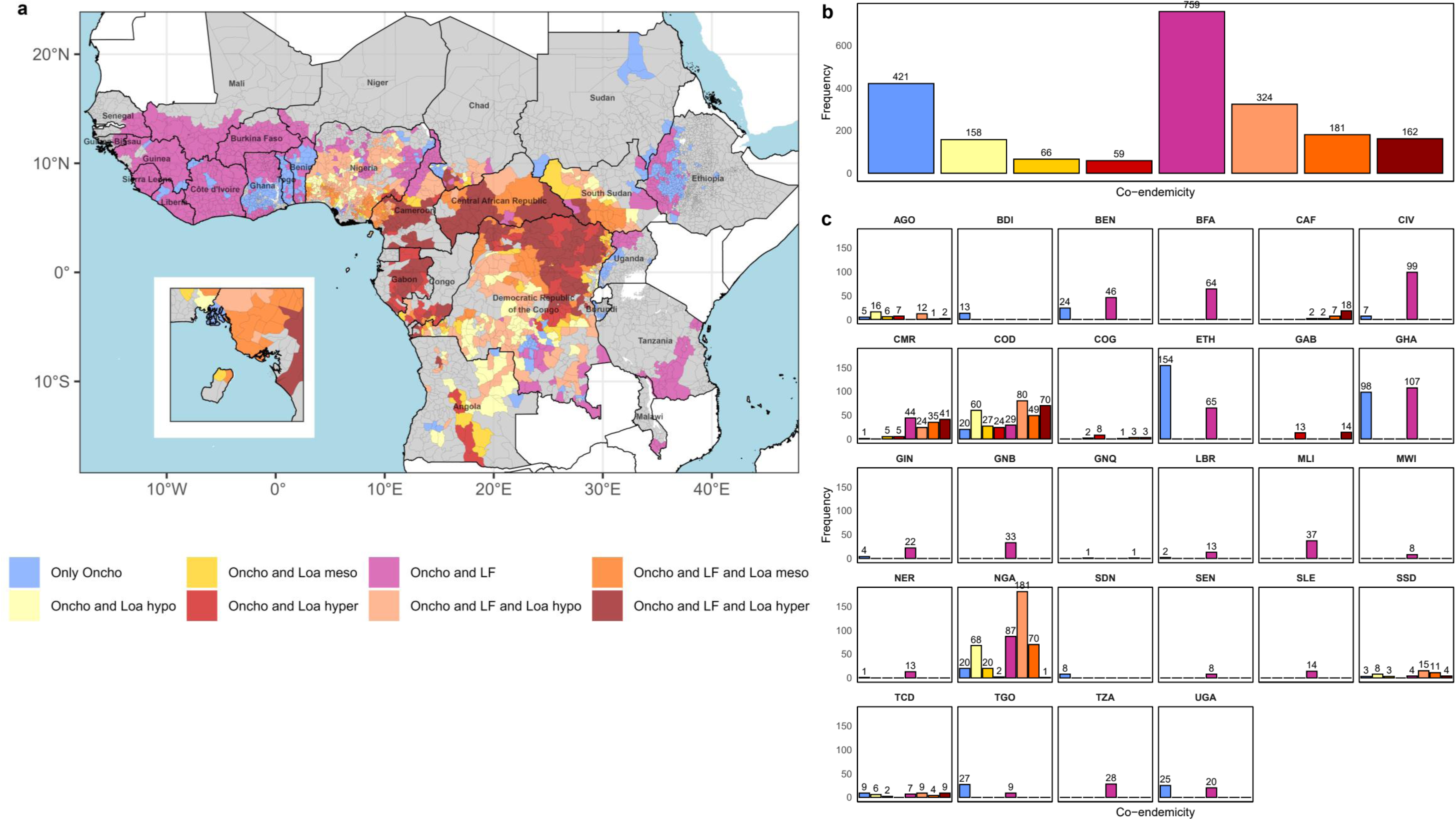
Co-endemicity of onchocerciasis (oncho) with lymphatic filariasis (LF) and loiasis (loa) by implementation unit (IU). Classifications of co-endemicity status are presented: (**a**) across a map of endemic countries, (**b**) as a frequency distribution across all IUs, and (**c**) as a frequency distribution of IUs within each country. IUs are indicated by grey borders. Inset map shows Bioko Island (Equatorial Guinea) in (**a**). IUs in grey are either non-endemic or with no intervention history (classified as ‘Unknown’ or ‘Not reported’ under the “Endemicity” variable in ESPEN)^22^. Loiasis endemicity classifications in the ESPEN loiasis databse^79^ are according to RAPLOA^80^. Country abbreviations are as in Fig. 5.

A total of 1,659 (77.9%) IUs are classified as ‘MDA continues’, 133 (6.2%) as ‘MDA stopped’, 309 (14.5%) as “some previous IVM; potentially requiring OEM and/or SM’ with 166 (53.7%) of these in former OCP areas (indicated by (*)), and 29 (1.3%) as ‘Treatment-naïve; potentially requiring OEM and/or SM’ as of 2022 (Fig. 8).

**Fig. 8.**
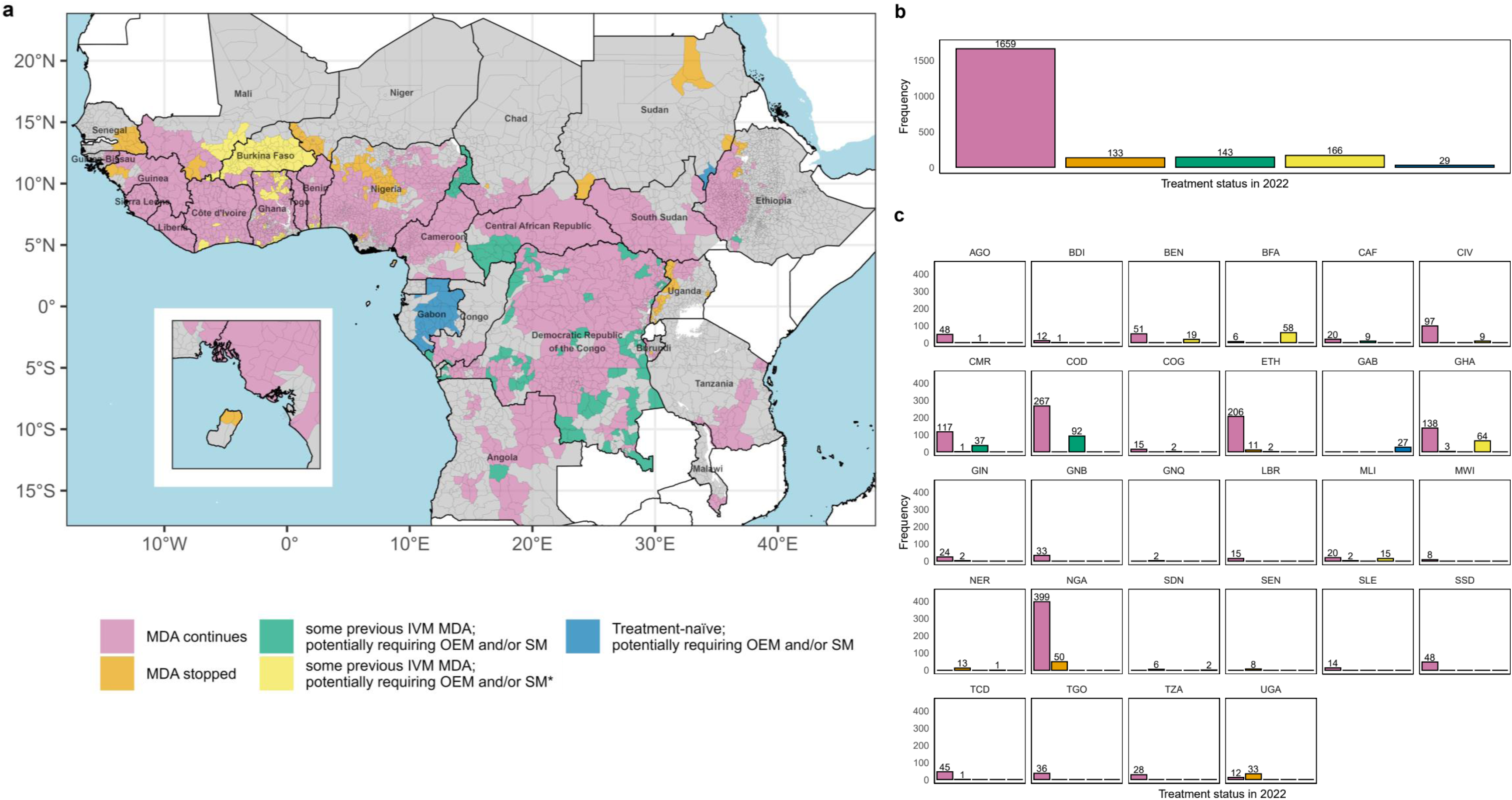
Ivermectin mass drug administration (MDA) treatment status in 2022. Treatment status in 2022 is presented: (**a**) across a map of endemic countries, (**b**) as a frequency distribution across all IUs, and (**c**) as a frequency distribution of IUs within each country. IUs are indicated by grey borders. Inset map shows Bioko Island (Equatorial Guinea) in (**a**). IUs in grey are either non-endemic or with no intervention history (classified as ‘Unknown’ or ‘Not reported’ under the “Endemicity” variable in ESPEN)^22^. Country abbreviations are as in Fig. 5. OEM: Onchocerciasis Elimination Mapping^72^; SM: Suitability Mapping^73^; (*): indicates IUs in former OCP countries potentially requiring OEM/SM.

The HISTONCHO database contains 34 variables for each IU. Table S1 in Supplementary File 1 lists the variable names, their descriptions and the values they can take (numerical or categorical), as well as their provenance (source).

## Technical Validation

Selection of information from a wide range of data and literature sources to inform the onchocerciasis intervention histories resulted in several challenges. The final compiled HISTONCHO database^84^ contains a mixture of detailed subnational information for a small number of countries, and coarse country- and regional-level information, particularly for the 1975-2012 period.

Where we did not have country-specific information on implementation of biannual MDA, we assumed that only annual MDA has been delivered. The “Cum_MDA” variable in the ESPEN database^22^ indicates how many previous MDA rounds have taken place. However, while this variable gives some indication at the IU level of the number of rounds prior to 2013-2014, it does not provide sufficient information to help backfill MDA characteristics for a given IU. For example, it gives no information on when an IU started MDA, in which years MDA did not occur, or when biannual MDA took place.

For former OCP countries, we used the OCP phase boundaries from the shape-files in O’Hanlon *et al*.^10^ to inform when interventions began in each IU, including both vector control and MDA. Literature sources (Table 1) indicated what interventions were implemented and their timings under each OCP phase, by country. Therefore, we simplified the intervention histories by applying this information across all IUs within a country and OCP phase, rather than using the “Cum-MDA” variable in the ESPEN database^22^.

For former APOC countries, the APOC report^26^ contains details on the year when MDA started for each country and provides coverage estimates. We used the APOC 2015 report^26^ to apply country-level coverages of total population (Table 2) to the IU level. We made the simplifying assumption that coverages are the same across IUs within a country (through to 2012), with MDA beginning at the same time in each IU within a country.

One variation that could be explored would be to produce an alternative intervention history dataset by using the “Cum_MDA” variable in ESPEN^22^. Even so, assumptions would still need to be made regarding coverage, whether MDA was continuous year-on-year, and its frequency. For former OCP countries in particular, information from the years preceding ESPEN on subnational total population coverage, especially in the earliest years of MDA implementation, would be essential to refine country-specific histories^53,54^.

We attempted to mitigate uncertainty in intervention characteristics prior to 2013 by working directly with implementation partners and Federal Ministries of Health (FMOH) whenever possible, or using other available literature sources. A collaboration with the Federal Ministry of Health and Social Welfare of Nigeria and implementation partners, such as The Carter Centre (TCC) and Sightsavers is an example of successful engagement at country level, which resulted in inclusion of detailed subnational MDA data. Information was supplied by the Nigeria FMOH directly^60^ on coverage values for each year, dating back to 1990 at the Local Government Area (LGA) level (Admin level 2). Information on biannual treatment was also supplied by the FMOH of Nigeria and annotated by TCC and Sightsavers^61^. We also received subnational information from the Republic of the Sudan FMOH and TCC to refine IU-level MDA information^65^. In the case of Uganda, we used a detailed review by Katabarwa *et al.*^69^, which documented when MDA took place and its frequency, as well as when vector control was implemented, and whether vector elimination (of *S. neavei*) was achieved in each focus. (IUs were mapped to these foci because district information was provided^69^.) For Togo, we used two papers^53,54^ that had analysed data shared by the Ministry of Health NTD Programme/National Onchocerciasis Elimination Programme to refine histories, with detailed information for SIZs at prefecture level. These examples of successful collaborations, or the availability of detailed literature to refine subnational information, are crucial to improve the validity of the HISTONCHO database^84^. It is paramount to build on these experiences to develop similar collaborations with other countries. For example, we have engaged with the Federal Democratic Republic of Ethiopia MOH through TCC, with the aim of refining timings/locations of biannual and quarterly (four times per year) MDA, the latter implemented to tackle persistent transmission in one subfocus^86^.

### Limitations

In the 2013-2022 period, we were able to include IU-level variation across each country by using the ESPEN database^22^. However, limitations associated with the ESPEN database still resulted in simplifications to the final intervention histories for this period. In 2013 for example, other than for specific countries such as Nigeria, Uganda and Sudan (Table 2), there were very limited data available. Therefore, we assumed that an IU should have MDA for 2013 when previous MDA was indicated by the “Cum_MDA” variable or by our histories through to 2012. However, we were not able to capture potential interruptions in 2013 based on this approach.

In HISTONCHO, MDA coverage is represented by the “Cov_Cat” variable, informed by the “EpiCov” variable in the ESPEN database^22^ and/or by “Cov_Raw” (Supplementary File 1, Table S1). “Cov_Cat” is a categorical variable that accounts for non-CDTI MDA, and for CDTI MDA to a maximum value of 65% of total population, as “EpiCov” values sometimes exceed 100%. This issue often arises in the ESPEN database where there is co-endemicity with LF (67% of IUs), as the population receiving treatment for LF is greater than the population requiring treatment for onchocerciasis. Unlike in LF—where the entire IU is targeted—the target units for onchocerciasis are smaller populations within the IU (due to its focal distribution), so the denominator is generally smaller. Coverages exceeding 100% in the ESPEN database can also occur when endemic countries upload coverage data on the ESPEN portal owing to their own resource constraints for regularly updating target population denominators, which results in older census estimates—which underestimate population growth and migration rates—being used, leading to inaccurate coverage data^87^. Additionally, in 309 (14.5%) of IUs, the classification of ‘some previous IVM; potentially requiring OEM and/or SM’ in the “Trt_Status_2022” variable reflects uncertainty in the MDA history of the IU and how this history is reflected in the current status of the IU. A total of 102 of these uncertain IUs are considered as ‘Unknown (under LF MDA)’ in the “Endemicity” variable of ESPEN^22^, and therefore, their MDA status could be due to treatment for LF rather than for onchocerciasis (yet they show a “Cum_MDA” value >0). Although the other 207 uncertain IUs are classified as either ‘Not reported’, ‘Unknown (consider Oncho Elimination Mapping)’, or ‘Non-endemic’, they also have a “Cum_MDA” value >0. To mitigate these uncertainties, we used the broad information in Table 1 and Table 2 when necessary. This would require further refinement through collaboration with ESPEN and onchocerciasis national control programmes.

The IU endemicity classifications in ESPEN are sometimes misaligned with the smooth prevalence predictions generated, at baseline, by the geostatistical approaches used by O’Hanlon *et al.*^10^ for OCP and Zouré *et al.*^23^ for APOC. ESPEN classifications would have been based on those in the OCP database and reported by APOC projects. Not only do these classifications change in these sources as prevalence decreases under intervention, but also they often correspond to river basins or transmission zones which may not directly translate into IU demarcations, especially as IU divisions have changed over time. Misalignment between endemicity classifications and MDA treatment information can also occur at country-level, as illustrated when comparing the Nigeria FMOH database^60^ with ESPEN’s, highlighting the challenges faced by the latter when centrally collating a large continental resource. Our work to link ancestral (‘original’) to subsequently subdivided (‘derived’) IUs attempted to address some of these issues by backfilling intervention histories; however, we still had to infer the endemicity status of the derived IUs and make the simplifying assumption that the MDA characteristics (whether MDA occurred and its coverage) for the larger original IU would apply to the smaller, resultant IUs in the same manner as before the split.

We excluded 494 IUs for which we deemed their treatment-naïve status as uncertain (Fig. 4). Although they have “Cum_MDA” = 0 in the ESPEN database, country-specific review—when this was possible—highlighted some potential discrepancies that would require engagement with the national programmes to be resolved. For instance, the aforementioned collaboration with the Sudan FMOH resulted in narrowing the number of treatment-naïve to two well-documented IUs^65^.

Working through these limitations presents opportunities to work closely with ESPEN and endemic countries—which upload their data to the ESPEN portal—to support improving and refining datasets. For example, information on biannual frequency will be an important inclusion in future ESPEN updates. Currently, we are also unable to document the reason for IUs having stopped MDA where we indicate this has occurred (Fig. 8); for instance, whether this is due to successful stop-MDA surveys and reaching WHO transmission interruption thresholds^88^, entering the PTS phase, or other programmatic considerations. Working with countries and ESPEN to ground-truth the ‘MDA stopped’ classification and the reasons for this would be an important enhancement to the HISTONCHO database^84^. HISTONCHO would also benefit from working together with ESPEN to host onchocerciasis epidemiological data to improve understanding of the impact of the interventions whose histories we have compiled.

## Usage Notes

The HISTONCHO database^84^ can be used by global health donors, country programmes, and implementation partners to understand the evolution of onchocerciasis control and elimination programmes, and by data analytics experts for data analysis and modelling of onchocerciasis control programmes at the IU level across SSA. The distribution of onchocerciasis-loiasis co-endemic areas (as well as that of these two filariases when co-endemic with LF) can also be helpful to inform where to deploy integrated mapping strategies to confirm the endemicity levels of the co-occurring filarial infections, and deliver treatment strategies that help circumvent or mitigate the problem of SAEs following treatment of highly microfilaraemic loiasis^76,89^. The distribution of onchocerciasis-LF co-endemic areas may also be used to decide where integrated transmission assessment surveys may be necessary^90^. This would require using HISTONCHO together with detailed spatial and temporal epidemiological data^53,54,91^. One such use case would be to simulate the impact of MDA programmes from the start of interventions within each IU using transmission dynamics models. Our “Cov_Cat” variable could be used and improved to better inform reliable total population coverage^87^, bearing in mind that robust data on treatment adherence will also be crucial^92^. The number of rounds (“Number_Rounds”) variable supports specifying annual or biannual treatment frequencies (and could adapted to reflect increased frequency^86^), and “Vector_Control” specifies whether anti-vectorial interventions have been implemented (by year, to account for their duration) and/or have eliminated a vector species^37,57,69^. It has been shown that information on baseline endemicity, number of years of treatment, biannual treatment frequency, treatment coverage, and vector elimination are significant determinants of onchocerciasis elimination prospects^6^.

## Code Availability

The code used to reconstruct the intervention histories, using the data sources described throughout the **Methods** section, is available at the following GitHub link: https://github.com/mrc-ide/HISTONCHO. Updates to the HISTONCHO database are encouraged through pull requests.

## Data Availability

All data produced are available online at https://zenodo.org/records/15390119

## Abbreviations

APOC: African Programme for Onchocerciasis Control
CDTI: Community-Directed Treatment with Ivermectin
ESPEN: Expanded Special Project for Elimination of Neglected Tropical Diseases
FMOH: Federal Ministry of Health
IA: Ivermectin plus Albendazole
IVM: ivermectin
JRF: Joint Reporting Form (of the WHO)
LF: lymphatic filariasis
LGA: local government area
MDA: mass drug administration
mf: microfilarial
mfp: microfilarial prevalence
NTD: neglected tropical disease
OEM: Onchocerciasis Elimination Mapping
OCP: Onchocerciasis Control Programme in West Africa
PTS: post-treatment surveillance
RAPLOA: Rapid Assessment Procedure for Loiasis
REMO: Rapid Epidemiological Mapping of Onchocerciasis
SAE: severe adverse event
SIZ: Special Intervention Zone
s.l.: sensu lato
SM: Suitability Mapping
SSA: sub-Saharan Africa
TCC: The Carter Center
WHO: World Health Organization.

## Acknowledgements

This work was supported by the Bill & Melinda Gates Foundation through the NTD Modelling Consortium (INV-030046). M.A.D. and M.-G.B. acknowledge funding from the MRC Centre for Global Infectious Disease Analysis (MR/X020258/1), funded by the UK Medical Research Council (MRC). This UK-funded award is carried out in the frame of the Global Health EDCTP3 Joint Undertaking. We wish to thank Dr Renata Retkute for her assistance in identifying annual and biannual treatment frequencies in Ethiopia, and Dr Mutono Nyamai for her help in identifying foci across sub-Saharan Africa for which elimination of onchocerciasis transmission has been reported.

## Author contributions

Conceptualisation: M.A.D., M.W., and M.-G.B. Data curation: M.A.D. Formal analysis: M.A.D., M.W., and A.R. Funding acquisition: M.W., and M.-G.B. Investigation: M.A.D., J.E.C., C.F. and M.-G.B. Methodology: M.A.D., M.W., and M.-G.B. Resources: J.E.C., E.G., G.S.N., A.T., E.M., A.M.A.A., J.C., P.B., R.B., W.A.S., and M.-G.B. Software: M.A.D., and A.R. Supervision: M.W., and M.-G.B. Validation: M.A.D., M.W., A.R. W.A.S., and M.-G.B. Visualisation: M.A.D. Writing – original draft: M.A.D., M.W., and M.-G.B. Writing – review & editing: M.A.D., M.W., A.R., J.E.C., E.G., G.S.N., A.T., E.M., A.M.A.A., J.C., P.B., C.F., R.B., W.A.S., and M.-G.B.

## Additional information

**Supplementary information** The online version contains supplementary materials.

**Correspondence** and requests for materials should be addressed to M.A.D. and M.G.B.

