## Supplementary Information File 1 for "HISTONCHO: A database of intervention histories for onchocerciasis control & elimination in sub-Saharan Africa"

### Title

**Table S1. HISTONCHO database variable names, descriptions and sources of information for the 34 variables included**

| Variable name | Description | Reference; Main text Table(s) & Figure(s) |
|---|---|---|
| IU_ID_MAPPING | ESPEN IU ID, updated to account for changes in Implementation Unit demarcations | ESPEN (2022)[1] |
| IUs_NAME_MAPPING | Name of Implementation Unit, updated to account for changes in Implementation Unit demarcations | ESPEN (2022)[1] |
| IU_CODE_MAPPING | ESPEN IU alphanumeric code (ISO3Code+ADMIN1+IU_ID), updated to account for changes in Implementation Unit demarcations | ESPEN (2022)[1] |
| ADMIN0ISO3 | Country ISO3 Code | ESPEN (2022)[1] |
| Endemicity | Reported Onchocerciasis Endemicity status through Joint Reporting Form (JRF) after final correction | ESPEN (2022)[1] |
| MAX_Endemicity | Defines the 'maximum' level of classification in the "Endemicity" for an IU across all ESPEN years (2013-2022) = i) 'Endemic' if this designation appears between 2013-2022, ii) 'Unknown', iii) 'Not reported', iv) 'Non-endemic' | – |
| Control_Prog | Indicates whether the IU is considered under the former Onchocerciasis Control Programme in West Africa or African Programme for Onchocerciasis Control = i) 'OCP', ii) 'APOC' | Table 1, Table 2 & Figure 1 |
| OCP_PHASE | Defines which OCP phase an IU belongs to (only former OCP countries) = i) 'Former OCP Phase I - 1975', ii) 'Former OCP Phase II - 1976', iii) 'Former OCP Phase III East - 1977', iv) 'Former OCP Phase III West - 1977', v) 'Former OCP Western Extension - 1989', vi) 'Former OCP Western Extension - 1990', vii) 'Former OCP Western Extension - 1990 (March)', viii) 'Former OCP Southern Extension - 1988', ix) 'Former OCP Southern Forest Extension-1990', x) 'Former OCP: Non-Control' | O'Hanlon *et al.*[2]; Figure 1 |
| SIZ_Label | Indicates whether the IU is within a former Special Intervention Zone (SIZ) for that year (only former OCP countries) = i) 'SIZ', ii) 'NA' (non-SIZ) | Table 1 & Figure 1 |
| Endemicity_Baseline | Onchocerciasis endemicity classification = i) 'Hypoendemic', ii) 'Mesoendemic', iii) 'Hyperendemic' based on the mean (point estimate) pre-control microfilarial prevalence for OCP countries or converted from Rapid Epidemiological Mapping of Onchocerciasis (REMO) nodule prevalence for APOC countries | O'Hanlon *et al.*[2]; Zouré *et al.*[3]; Coffeng *et al.*[4]; Coffeng *et al.*[5] |
| Year | Year | ESPEN (2022)[1] |
| PopTot | Reported/estimated total population for 2013-2022 | ESPEN (2022)[1] |
| PopPreSAC | Reported/estimated pre-school-age children population for 2013-2022 | ESPEN (2022)[1] |
| PopSAC | Reported/estimated school-age children population for 2013-2022 | ESPEN (2022)[1] |
| PopAdult | Reported/estimated adult population for 2013-2022 | ESPEN (2022)[1] |
| PopReq | Reported total population requiring treatment (JRF) for 2013-2022. JRF: Joint Reporting Form | ESPEN (2022)[1] |
| PopTrg | Total population targeted for treatment for 2013-2022 | ESPEN (2022)[1] |
| PopTreat | Total population treated for 2013-2022 | ESPEN (2022)[1] |
| MDA_Scheme | Mass drug administration (MDA) scheme implemented (which provides maximum coverage across different schemes) including values for ivermectin (IVM) = i) 'IVM'; ii) albendazole-ivermectin | ESPEN (2022)[1] (for 2013–2022); APOC (2015)[6] (for 1975- |

| | | |
|---|---|---|
| | = 'ALB-IVM'; iii) 'Not delivered'; iv) 'Pre-ESPEN IVM' for 1975-2012'; v) Not applicable ='NA'* | 2012); Table 1 & Table 2 |
| Cum_MDA | Cumulative MDA rounds since the inception of MDA interventions with ivermectin | ESPEN (2022)[1] |
| Cov | Programme coverage for total population (total treated/total pop targeted for treatment) 2013-2022, expressed as a proportion | ESPEN (2022)[1] |
| EpiCov | Epidemiological coverage for total population (total treated/total pop requiring treatment) for years 2013-2022, expressed as a proportion | ESPEN (2022)[1] |
| Vector_Control | Indicates the presence of vector control = i) '1' (present), ii) '2' (vector elimination or disappearance), iii) '0' (no vector control) | Table 1 & Table 2 |
| Non-CDTI_MDA | Indicates annual MDA before Community-Directed Treatment with Ivermectin (CDTI) = i) '1' (annual MDA), ii) '0' (no MDA) | Table 1 & Table 2 |
| CDTI_MDA | Indicates MDA once CDTI commences = i) '1' (CDTI MDA), ii) '0' (no CDTI MDA) | APOC (2015)[6]; Table 1 & Table 2 |
| Biannual_CDTI_MDA | Indicates biannual CDTI MDA = i) '1' (biannual MDA), ii) '0' (no biannual MDA) | Table 1 & Table 2 |
| Number_Rounds | Indicates the number of yearly MDA rounds = i) '1', ii) '2' depending on frequency (sum of "Non-CDTI_MDA", "CDTI_MDA" and "Biannual_CDTI_MDA") | – |
| Cov_Raw | Raw coverage values (continuous numerical variable), expressed as a proportion | ESPEN (2022)[1]; APOC (2015)[6]; Table 2 |
| Cov_Source | Indicates the source of the coverage data informing "Cov_Raw" | ESPEN (2022)[1]; APOC (2015)[6]; Table 2 |
| Cov_Cat | Categorised coverage value informed by "EpiCov" and/or "Cov_Raw" = i) '0.52' for "Non-CDTI_MDA" = '1'; ii) '0.25' for "CDTI_MDA" = '1' when "Epi_Cov" and/or "Cov_Raw" is >0 but <0.65, iii) '0.65' for "CDTI-MDA" =1 when "Epi_Cov" and/or "Cov_Raw" ≥0.65 | – |
| MDA_VC_Mapping | Summarises the type of intervention history for the IU (across all years) = i) 'Annual MDA' where only annual ivermectin MDA has been implemented; ii) 'Biannual MDA' if there has been any biannual MDA, iii) 'Annual MDA and vector control', iv) 'Biannual MDA and vector control', v) 'Treatment-naïve' (IUs without any intervention) | Figure 5 |
| Cum_MDA_Rev | Revised cumulative number of MDA rounds based on back-dated histories (calculated using "Number_rounds") | Figure 6 |
| Co_endemicity | Indicates co-endemicity status with loiasis and/or lymphatic filariasis (LF) = i) 'Only Oncho', ii) 'Oncho and LF', iii) 'Oncho and Loa hypo' (hypoendemic loiasis), iv) 'Oncho and Loa meso' (mesoendemic loiasis), v) 'Oncho and Loa hyper' (hyperendemic loiasis), vi) 'Oncho and LF and Loa hypo', vii) 'Oncho and LF and Loa meso', viii) 'Oncho and LF and Loa hyper'. (For Oncho pre-control endemicity see "Endemicity_Baseline") | Figure 7 |
| Trt_Status_2022 | Indicates status of the IU in 2022 = i) 'MDA continues', ii) 'MDA stopped', iii) 'some previous IVM; potentially requiring OEM and/or SM', iv) 'some previous IVM; potentially requiring OEM and/or SM*', v) 'Treatment-naïve; potentially requiring onchocerciasis elimination mapping (OEM) and/or suitability mapping (SM)' (*) indicates in former OCP areas | Figure 2 & Figure 8 |

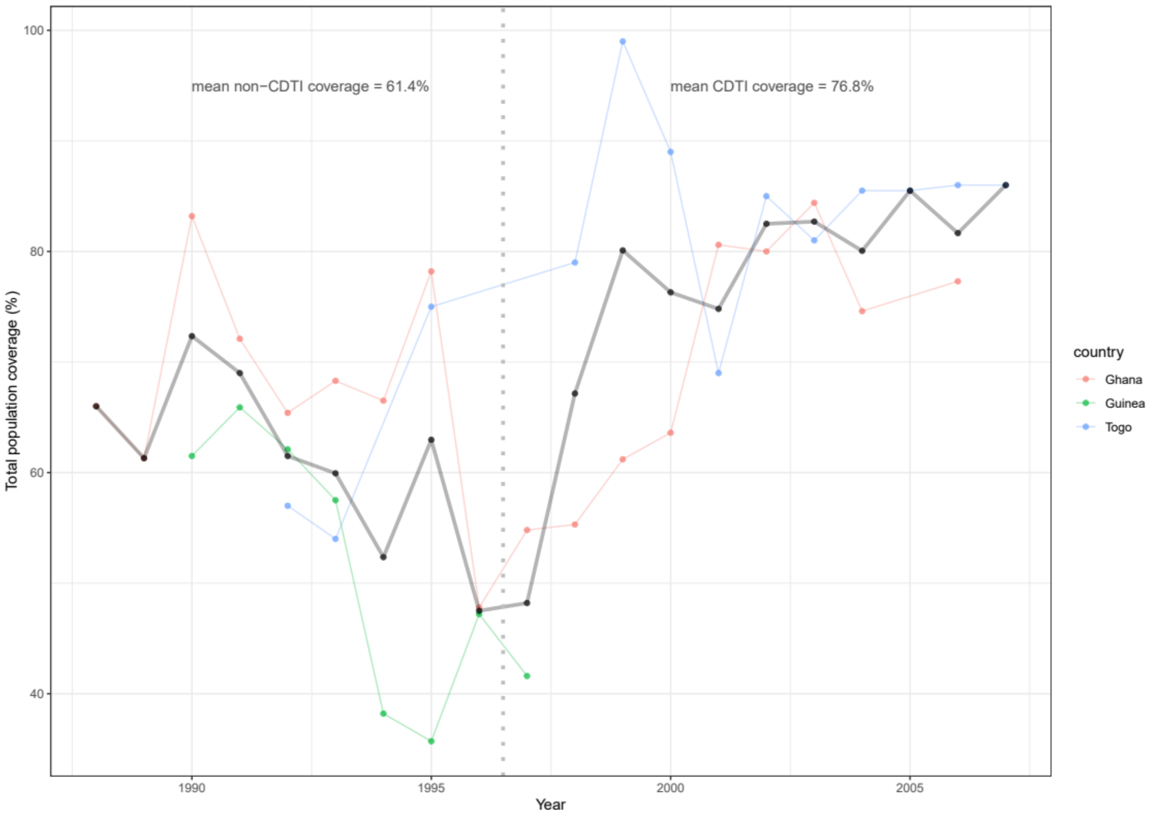


**Fig. S1 Coverage temporal trends for three indicator villages within Special Intervention Zones (SIZ) of the former Onchocerciasis Control Programme in West Africa (OCP).** The villages are: Asubende (Ghana, red), Serekoroba (Guinea, green) and Titira (Togo, blue). Coverage of total population is plotted for each year during the period 1988–1996, prior to the adoption of Community-Directed Treatment with Ivermectin (non-CDTI) and during 1997–2007, subsequent to CDTI implementation. The thick grey line is the mean coverage for each year. The dashed vertical light grey line signals the switch in ivermectin distribution from non-CDTI to CDTI. On average, during the non-CDTI period the coverage was 61.4% in contrast to the average coverage of 76.8% for the CDTI period, suggesting that CDTI led to an increase in coverage of approximately 25% in these examples[7].
